# Rapid response flow cytometric assay for the detection of antibody responses to SARS-CoV-2

**DOI:** 10.1101/2020.05.09.20091447

**Authors:** Dennis Lapuente, Clara Maier, Pascal Irrgang, Julian Hübner, Sophia Peter, Markus Hoffmann, Armin Ensser, Katharina Ziegler, Thomas H. Winkler, Torsten Birkholz, Andreas E. Kremer, Philipp Steininger, Klaus Korn, Frank Neipel, Klaus Überla, Matthias Tenbusch

**Affiliations:** Institute of Clinical and Molecular Virology, University Hospital Erlangen, Friedrich-Alexander University Erlangen-Nürnberg, 91054 Erlangen, Germany; Infection Biology Unit, German Primate Center - Leibniz Institute for Primate Research, 37077 Göttingen, Germany; Institute of Clinical Hygiene, Medical Microbiology and Infectiology, Paracelsus Medical University, Nürnberg, Germany; Department of Biology, Division of Genetics, Nikolaus-Fiebiger-Center for Molecular Medicine, Friedrich-Alexander University Erlangen-Nürnberg, Erlangen, Germany; Department of Anaesthesiology, University Hospital Erlangen, Erlangen, Germany; Department of Medicine 1, Gastroenterology, Pneumology and Endocrinology, University Hospital Erlangen, Friedrich-Alexander University Erlangen-Nürnberg, 91054 Erlangen, Germany

## Abstract

SARS-CoV-2 has emerged as a previously unknown zoonotic coronavirus that spread worldwide causing a serious pandemic. While reliable nucleic acid-based diagnostic assays were rapidly available, there exists only a limited number of validated serological assays. Here, we evaluated a novel flow cytometric approach based on antigen-expressing HEK 293T cells to assess spike-specific IgG and IgM antibody responses. Analyses of 201 pre-COVID-19 sera proved a high assay specificity in comparison to commercially available CLIA and ELISA systems, while also revealing the highest sensitivity in specimens from PCR-confirmed SARS-CoV-2 infected patients. Additionally, a soluble Angiotensin-Converting-Enzyme 2 (ACE-2) variant was established as external standard to quantify spike-specific antibody responses on different assay platforms. In conclusion, our newly established flow cytometric assay allows sensitive and quantitative detection of SARS-CoV-2-specific antibodies, which can be easily adopted in different laboratories and does not rely on external supply of assay kits.

## Introduction

In early December 2019, a novel zoonotic coronavirus (CoV) caused a cluster of pneumonia cases in Wuhan, China (1). Since then, the virus has spread globally and caused a pandemic with over 3,435,000 confirmed infections and about 240,000 fatalities (as of May 4^th^ 2020) (2). Due to its phylogenetic similarity to the Severe Acute Respiratory Syndrome Related Coronavirus (SARS-CoV-1), the novel CoV was named SARS-CoV-2 (3). The acute respiratory disease induced by SARS-CoV-2 is called coronavirus disease 19 (COVID-19).

The identification of acutely infected individuals by the detection of viral RNA by real-time PCR (4) was implemented rapidly in the health care of most countries. While this method is highly valuable for the diagnosis of acute COVID-19 cases, specific serological methods are urgently needed to determine seroconversion in general and more specifically to characterize the humoral response against SARS-CoV-2. Robust, validated serological approaches are essential to track transmission events in individuals that have already cleared the infection especially after mild or symptom-free disease. With increasing numbers of immune individuals, serological tests will also help to understand epidemiological aspects of the pandemic and to employ SARS-CoV-2 immune staff in critical frontline positions at hospitals or nursing homes. In addition, validated serological methods are essential to evaluate novel vaccine candidates in clinical studies.

Together with the 2003 SARS-CoV-1 and the 2012 Middle East Respiratory Syndrome Coronavirus (MERS-CoV) epidemic, the SARS-CoV-2 pandemic represents the third betacoronavirus in twenty years that crossed the species barrier and resulted in a significant number of human infections. At the same time, four other CoVs are endemic in the human population (two alphacoronaviruses: CoV-NL63 and -229E, two betacoronaviruses: CoV-OC43 and -HKU1) that cause episodes of common cold in humans in all parts of the world (5). CoVs are enveloped single-stranded RNA viruses that contain four structural proteins: membrane (M), envelope (E), spike (S), and nucleocapsid (N). From SARS-CoV-1 it is known that N and S proteins are the most immunogenic viral antigens, while only S-specific antibodies can mediate virus neutralization (6,7). Therefore, N- and S-specific antibody responses should be first choice parameters for a sensitive serology (8). However, depending on the study cohort, up to 90% of the population is seropositive for common cold CoVs (9-11). Thus, a careful validation of the assay specificity is required in CoV serology.

Here, we describe a novel flow cytometric assay to determine SARS-CoV-2 spike protein-specific antibodies in serum samples. The virus-free assay relies on reagents and devices that are available in many medical and biological research labs and therefore can be easily adopted in a decentral manner without the need for commercial kits or products that are prone to shortage.

## Materials and methods

### Serum samples

Anonymized, random sera (n=180) were selected from the sample repository of the diagnostics department of the Institute for Clinical and Molecular Virology at the University Hospital Erlangen to evaluate the specificity of the novel diagnostic test. Samples were collected until August 2019 (further denominated as pre-COVID-19 era) and no longer needed for diagnostic purposes and assigned for disposal. Those specimens were not characterized in regard to anti-HCoV antibody status. 21 sera from eight patients with PCR-confirmed endemic HCoV infections were additionally included. These samples were collected at least one week before and two to four weeks after HCoV infection. These include 4x HKU-1, 2x 229E, 1x NL63, 1x OC43 infections. Post-infection sera were sampled twice from some patients (Table 1). Additionally, 60 specimens from 34 individuals with a PCR-confirmed SARS-CoV-2 infection (some sampled longitudinally) were obtained. The majority is derived from a newly established biobank for COVID-19 patients at the University Hospital Erlangen. The data are collected in accordance with ethical requirements. The implementation of the biobank has been approved by the local ethics committee of the UK Erlangen under the licence number AZ. 174_20 B. Five out of 60 were derived from plasma donors after (patients’ informed consents; approved by local ethics committee of the FAU; AZ. 2020, 49_20B). Another set of sera was collected from thirteen COVID-19 patients at the Hospital Nürnberg Nord at different time points after the PCR-confirmation (Table 2). All sera were sampled for recent diagnostic purpose and have been tested for seroconversion in the EuroImmun ELISA at the Institute of Clinical Hygiene, Medical Microbiology and Infectiology, Paracelsus Medical University, Hospital Nürnberg, Germany. All clinical specimen were used in anonymous form for retrospective analyses.

**Table 1.** Validation of the flow cytometric assay for SARS-CoV-2-specific IgM and IgG with serum samples collected before the COVID-19 outbreak and a set of sera from PCR- confirmed SARS-CoV-2 infections

|  | Samples<br>(# of patients) | IgM+ (%)* | IgM- (%)* | IgG+ (%) | IgG- (%) |
| --- | --- | --- | --- | --- | --- |
| Pre-COVID-19 sera | 180 (n.d.) | 0 (0) | 84 (100) | 2 (1.1) | 178 (98.8) |
| HCoV+ (endemic, pre) | 8 (8) | 0 (0) | 8 (100) | 0 (0) | 8 (100) |
| HCoV+ (endemic, post) | 13 (8) | 0 (0) | 13 (100) | 0 (0) | 13 (100) |
| Recent SARS-CoV-2 PCR+ | 60 (34) | 48 (87.3) | 7 (12.7) | 60 (100) | 0 (0) |

n.d., not determined. \*, IgM testing was not available for initial tests, which explains the lower sample number

**Table 2:** Analysis of serum samples from COVID-19-infected individuals at various time points relative to PCR-confirmation by EuroImmun ELISA, in-house flow cytometric assay, and Yhlo CLIA

| Sample | Sampling<br>relative to<br>PCR [days] | EuroImmun ELISA |  |  |  | Flow cytometry assay |  |  |  | Yhlo CLIA |  |  |  |
| --- | --- | --- | --- | --- | --- | --- | --- | --- | --- | --- | --- | --- | --- |
|  |  | IgA | IgA<br>Ratio | IgG | IgG<br>Ratio | IgM | IgM<br>Ratio | IgG | IgG<br>Ratio | IgM | AU/ml | IgG | AU/ml |
| 1 | 0 | - | 0.4 | - | 0.2 | - | 1.9 | - | 3.1 | - | 0.54 | - | 0.52 |
| 2 | 0 | - | 0.1 | - | 0.2 | - | 1.2 | - | 0.95 | - | 1.18 | - | 2.29 |
| 3 | 0 | + | 1.9 | +/- | 0.9 | + | 9 | + | 24.1 | - | 5.46 | - | 1.72 |
| 4 | 0 | + | 3 | + | 2.7 | + | 17 | + | 90.7 | + | 362.64 | + | 30.94 |
| 5 | +2 | + | 4.5 | + | 4.5 | + | 22.2 | + | 160 | + | 104.09 | + | 49.6 |
| 6 | +5 | + | ≥11 | + | 9.4 | + | 41 | + | 112.6 | + | 254.86 | + | 61.55 |
| 7 | +6 | + | ≥11 | + | 9.4 | + | 42.7 | + | 166.3 | + | 605.83 | + | 47.87 |
| 8 | +7 | + | ≥11 | + | 14.5 | + | 15.4 | + | 95.9 | + | 15.47 | + | 112.52 |
| 9 | +8 | + | ≥11 | + | 12.6 | + | 56 | + | 263.3 | + | 246.58 | + | 101.62 |
| 10 | +8 | + | 4.8 | +/- | 0.8 | + | 31 | + | 103.4 | - | 6.48 | + | 16.94 |
| 11 | +9 | + | ≥11 | + | 15.7 | + | 52 | + | 321.8 | + | 106.43 | + | 161.04 |
| 12 | +12 | + | 10.1 | + | 5.6 | + | 48.3 | + | 345.5 | + | 43.27 | + | 186.03 |
| 13 | +14 | + | ≥11 | + | 14 | + | 37.1 | + | 261.5 | - | 7.76 | + | 155.16 |
| Reactivity |  | 11/13 |  | 9/13 |  | 11/13 |  | 11/13 |  | 8/13 |  | 10/13 |  |

\* Time gap between SARS-CoV-2 diagnosis by PCR and collection of blood sample. AU, arbitrary units. +, seropositive. -, seronegative. +/-, borderline. Ratios for IgG and IgA ELISA indicate the ratio between sample and calibrator. Ratios for the flow cytometric assay indicate the ratio between mock MFI and SARS-CoV-2 MFI for a given sample. Red/orange background indicates negative/borderline result.

### DNA plasmids

The pCG1_CoV_2019-S plasmid encoding the codon-optimized sequence of the SARS-CoV- 2 S protein was generated as described elsewhere (12). The plasmid pcDNA3.1 (Invitrogen) was used in the mock transfection control. Blue fluorescent protein (BFP) and red fluorescent protein-encoding (dsRed; from *Discosoma sp.)* plasmids were used as marker proteins for transfected 293T cells.

### Flow cytometric antibody assay

Human embryonic kidney cells (HEK 293T cells; ECACC 12022001) were maintained in Dulbecco’s modified Eagle’s medium (DMEM; Gibco, Cat #11960-044) containing 10% fetal calf serum (Capricorn Scientific, Cat #FBS-12A), 1% GlutaMAX (Gibco, Cat #35050-038), and 1% Penicillin/Streptomycin (Gibco, Cat #15140-122) at 37°C and 5% CO_2_. For the assay, 1.12×10^7^ cells were plated out (25 ml medium; 175 cm^2^ cell culture flask) and, 12-24 hours later, were transfected with 30 (μg pCG1_CoV_2019-S plus 15 μg fluorescent protein (BFP) by standard polyethylenimine transfection (3.5 ml DMEM, 67.5 μg polyethylenimine). As an internal control, a mock transfection was used with 30μg pcDNA3.1 and 15μg fluorescent protein (dsRed). 48 hours after the transfection, cells were harvested, resuspended in freeze medium (75% FCS, 10% DMSO, 3% Glucose in DMEM), and stored in 1 ml aliquots of 1×10^7^ cells at −80°C.

For the assay, aliquots of cells were thawed, washed once with PBS, and then resuspended in FACS buffer (PBS with 0.5% bovine serum albumin and 1 nmol sodium azide). 0.5×10^5^ cells of each of the two cell preparations (S- and mock-transfected) were seeded out per sample in a 96-well U-bottom plate. Serial dilutions of the standards or serum samples (1:100) were diluted in 100 μl FACS buffer and given on the cells (30 min, 4°C). 100 μl FACS buffer was added, cells were centrifuged (500 xg, 4°C, 3min; used for all following centrifugation steps), washed two times with 180 μl FACS buffer, and bound antibodies were stained with secondary detection antibodies diluted 1:300 in 100 μl FACS buffer (30 min, 4°C, anti-IgG- AF647, clone HP6017, Biolegend, Cat #409320; anti-IgM-BV711, clone MHM-88, Biolegend, Cat #314540). 100 μl PBS was added, cells were centrifuged, washed two times with 180 μl PBS, and fixed in 200 μl 2% paraformaldehyde in PBS (15 min, 4°C). Cells were centrifuged and washed once in 180 μl FACS buffer, before resuspended in 200 μl FACS buffer for flow cytometric analysis. Data were acquired on a BD LSRII or Thermo Fisher Attune Nxt cytometer and analysis was performed with FlowJo (Tree Star Inc.) or Flowlogic (Inivai Technologies).

### ACE-2-Fc standard

A PCR fragment containing the sequence coding for the extracellular domain of human ACE-2 lacking the secretory signal peptide (NM_021804.3, nucleotides 358 – 2520) fused at the 3’ end with a PCR fragment coding for the Fc-part of human IgG1 and a C-terminal myc/his tag was cloned into the expression vector pCEP4 (Thermo Fisher Scientific). The signal peptide of the murine IgG kappa-chain V-J2 was used instead of the ACE-2 signal peptide. The synthetic intron from pIRES (IVS, Takara Bio) was cloned via NheI restriction sites between the transcription start and the translation start site. Expression and purification of the Fc-fusion protein was done as described before (13). Briefly, HEK 293T cells were transfected by calcium phosphate method and kept in culture for six days. Cell culture supernatant was then harvested and cell debris removed by centrifugation. The pH of the supernatant was adjusted to 8.0 with NaOH and sterile filtered. The supernatant was then applied to a HiTrap Protein A HP column (GE Healthcare Life Sciences). ACE-2-Fc fusion protein was eluted by a pH step gradient using 0.1 M citrate buffer. ACE-2 Fc fusion protein eluted at pH 4.0 and the pH was immediately neutralized by the addition of 1M Tris buffer (pH 9).

As an external standard for IgG quantitation, a two-fold dilution series starting with 10 μg/ml of ACE-2-Fc was measured in the flow cytometric assay as described above. With this standard, we quantified the amount of ACE-2-binding equivalents in a plasma sample available in larger volume. Adjusting for molecular weight differences between ACE2-Fc and IgG, the anti-SARS-CoV-2 S IgG concentration in this plasma sample was determined.

### Enzyme-linked Immunosorbent Assay (ELISA)

Commercially available ELISA for the detection of anti-SARS-CoV-2 IgG (anti-S1-specific, EuroImmun, Cat #EI 2606-9601 G) and IgA (EuroImmun, Cat #EI 2606-9601 A) were performed according to the manufacturer’s protocols. Sera were diluted 1:101 (10μl sample + 1000μl sample buffer) and the optical density was detected at 450 nm at a multilabel plate reader (Victor X5, Perkin Elmer). A cut-off for a positive result was according to the manufacturer defined as a ratio of >1.1 between the specific specimen and the calibrator. Values between 0.8 and 1.1 were defined as “borderline”.

### Chemiluminescent Immunoassay (CLIA)

Commercially available magnetic bead-based CLIA for the detection of IgG (N- and S-specific, Shenzhen Yhlo Biotech, iFlash-SARS-CoV-2, Cat #C86095G) and IgM (Shenzhen Yhlo Biotech, iFlash-SARS-CoV-2, Cat #C86095M) were performed on a fully automated iFlash Immunoassay Analyzer (Shenzhen Yhlo Biotech). The assays were performed according to the manufacturer’s protocols. The IgG and IgM titer were automatically calculated as arbitrary units (AU/ml) and the cut-off value for a positive test was 10 AU/ml.

## Results

### Assay specificity for SARS-CoV-2 immune sera

The novel serological assay we evaluate here exploits 293T cells that express full-length SARS-CoV-2 spike protein in its natural conformation to bind antigen-specific IgM and IgG from patient sera with a subsequent quantification by secondary detection antibodies. In a multiplex approach with two populations each co-expressing a specific fluorescent protein (dsRed or BFP), non-antigen-expressing cells provide an internal specificity control. By this, one can control for unspecific binding of antibodies to cellular components leading to potentially false-positive results for example in patients with autoimmune diseases.

Figure 1 illustrates the gating strategy and the respective IgM and IgG mean fluorescence intensity (MFI) signals for three negative controls and three SARS-CoV-2 convalescent sera. None of the negative control sera S4-S6 led to a significant MFI increase in the S-expressing population compared to the mock control cells. In contrast, both for IgM and IgG, the MFI in the S-expressing cells were clearly increased compared to the mock cells indicating a specific binding of S-specific antibodies. Subsequently, we defined two cut-off criteria for a positive serological result: (i) the MFI of the test sample must be at least three-fold higher compared to the mean of three negative sera tested in parallel and (ii) the ratio of MFI SARS-CoV-2/MFI mock must be higher than 3.

**Figure 1.**
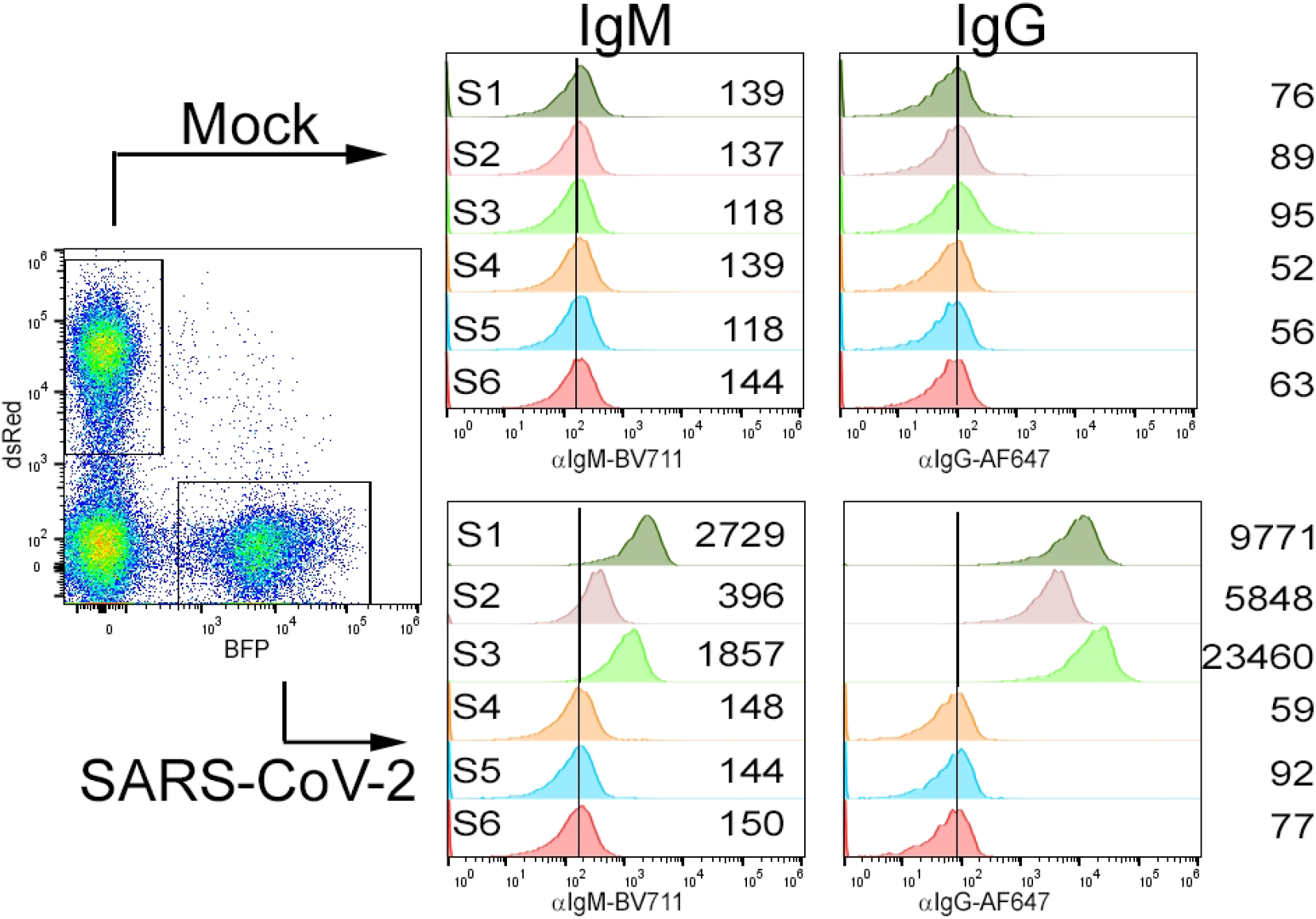
Representative samples measured in the flow cytometric assay. As described in the methods section, 293T cells expressing SARS-CoV-2 S protein and pcDNA3.1-transfected cells (mock) were incubated with COVID-19 patient sera (S1-S3) or with negative control sera (S4-S6). Bound IgM and IgG were detected with secondary detection antibodies. The left plot shows the gating of the target populations considering the co-transfected fluorescent proteins as transfection markers (BFP and dsRed). The right histograms depict IgM and IgG fluorescence signals in each sample for both cell populations, respectively. The mean fluorescence intensity is shown in numbers.

We evaluated the assay specificity and sensitivity with a set of 180 historic diagnostic samples that had not been analysed for antibody responses to endemic HCoV, with 21 sera derived from eight patients with confirmed endemic HCoV infections (pre- and post-infection sera sampled), as well as with 60 sera from SARS-CoV-2 PCR-positive individuals (Table 1). The results for IgM and IgG (MFI SARS-CoV-2/MFI mock ratios) are shown for a representative set of specimen in figure 2 and a summary of all sera tested for S-specific antibody responses is provided in table 1. With the cut-off criteria defined above, 60 out of 60 specimen collected from PCR-confirmed SARS-CoV-2 patients were IgG-positive (100%) and 48 out of 55 were tested IgM-positive (84.2%; not all samples characterized for IgM). Of note, the negative testing for IgM occurred in serum samples, which were still IgG positive and sampled most probably in the later stage of convalescence. Regarding the specificity of the assay, IgM exceeded the cut-off criteria only in patients with a previously PCR-confirmed SARS-CoV-2 infection, while none of the sera from uninfected individuals did so (0/105). For IgG, two out of 180 sera (1.1 %) sampled in the pre-COVID-19 era without any information about HCoV status surpassed the criteria for seroconversion. Importantly, none of the sera with a PCR-confirmed endemic HCoV infection (sampled 2-4wks post-infection) showed any IgG or IgM cross-reactivity, thus, indicating a high degree of assay specificity for SARS-CoV-2 seroconversion.

**Figure 2.**
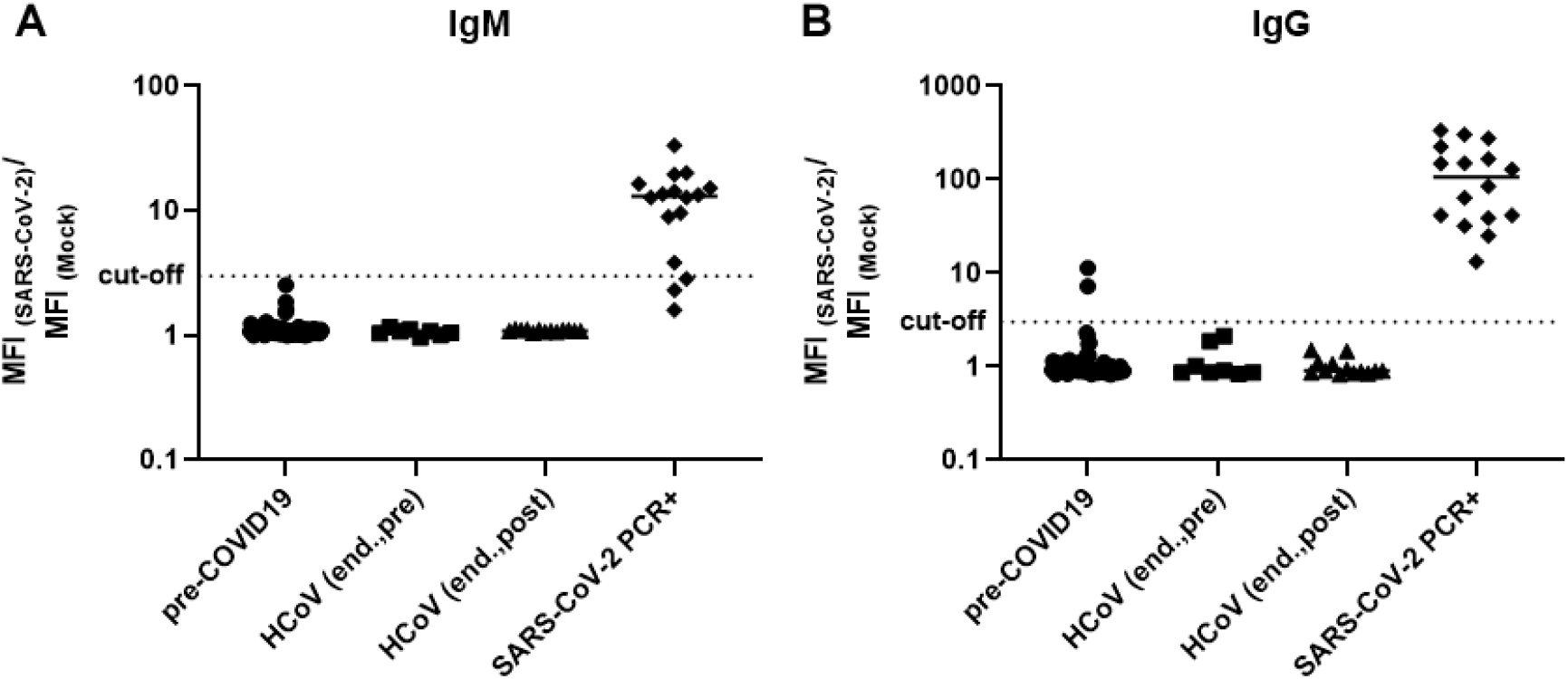
Analysis of SARS-CoV-2-specific IgM and IgG in serum samples from uninfected individuals or COVID-19 patients. The flow cytometric serological assay was performed with samples from the pre-COVID-19 era (n=82), samples from individuals with known endemic HCoV infection (n=8 before infection, HCoV (end., pre); n=13 after infection, HCoV (end., post)), and samples from PCR-positive COVID-19 patients (n=16). Shown are the ratios of the MFI values for SARS-CoV-2-expressing and mock-transfected cells for IgM (a) and IgG (b). The cut-off is depicted as dotted line and represents a ratio of 3. Shown are individual serum samples and the group mean. MFI, mean fluorescence intensity. end, endemic.

A longitudinal analysis of a patient starting at the day of PCR-confirmed SARS-CoV-2 infection (3^th^ April, day 0) presented specific seroconversion for IgG around day 8 (11^th^ April) and for IgM around day 10 (13^th^ April) although showing elevated levels of IgM below the cut-off already earlier (Fig. 3A). A second patient presented earlier IgM (day 3, 30^th^ March) than IgG seroconversion (day 7; Fig. 3B), but it is important to note that the exact infection events are unknown in both cases.

**Figure 3.**
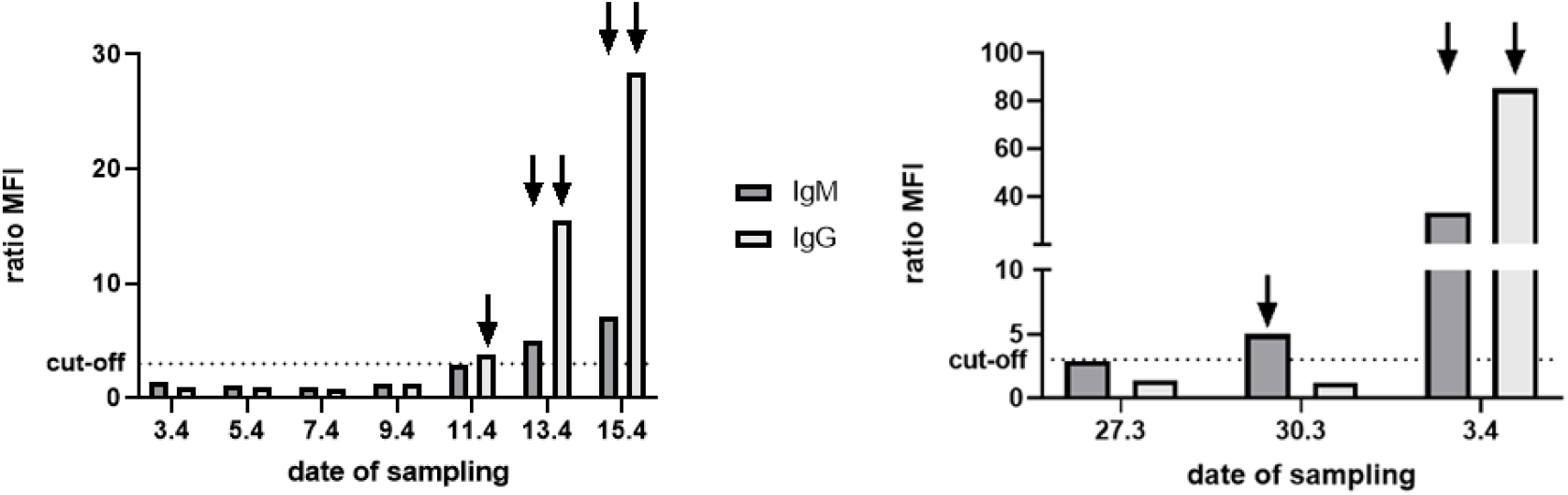
Longitudinal analysis of SARS-CoV-2 seroconversion in two COVID-19 cases. At each depicted date, serum samples were collected from the respective patient and analysed by the flow cytometric assay for IgM and IgG. The cut-off is depicted as dotted line and represents a ratio of the MFI values for SARS-CoV-2-expressing and mock-transfected cells of 3. Arrows indicate values above the cut-off.

### Performance compared to commercial kits

We further assessed thirteen serum samples from PCR-confirmed cases of SARS-CoV-2 infections in a comparative analysis with our flow cytometric assay, a commercial ELISA for IgA/IgG (spike subunit S1-specific; EuroImmun), and a commercial CLIA (N- and S-specific; Shenzhen Yhlo Biotech). Among those thirteen samples, eight were positive for SARS-CoV- 2-specific antibodies in all assays, while two specimens were uniformly negative (Table 2). Of note, those two sera were sampled at the day of PCR-confirmation, thus, seroconversion might not yet have been occurred. Similarly, another specimen (sample 3) was sampled at the same day as the first positive PCR test and showed IgG/IgM seroconversion in the flow cytometric assay, but did not show reactivity in the CLIA and only borderline reactivity in the IgG ELISA (“borderline” as defined by manufacturer). The flow cytometric serological assay for IgG and IgM as well as the ELISA for IgA showed the highest sensitivity with 11/13 specimens above the cut-off. Two sera were borderline positive in the IgG ELISA, while both positive in the flow cytometric assay and one positive in the IgG CLIA. Overall, this demonstrates a high sensitivity of our cytometric antibody detection for seroconversion in a direct comparison to commercially available detection kits. The lower detection limit was also confirmed by serial dilutions of selected positive samples. While 1-10,000 dilutions were still measured as seropositive by our flow cytometric assay, the two other kits revealed a negative result suggesting a higher analytical sensitivity of the flow cytometric test (data not shown).

### ACE-2-Fc as external standard for absolute quantification of samples

In order to allow quantitation of antibody responses, we developed an external standard based on the soluble SARS-CoV-2 entry receptor ACE-2 (12,14) fused to a human IgG fragment crystallizable region (Fc region). As depicted in figure 4A, the ACE-2-Fc standard binds to SARS-CoV-2-expressing HEK 293T cells in a concentration-dependent manner with a linear incline before a saturation phase at higher concentrations. The MFIs of the standard curve demonstrate a strong reproducibility with low inter-assay variation (Fig. 4A) and allow absolute quantitation of in-house standard sera or plasma. The anti SARS-CoV-2 IgG concentration in our standard plasma was determined using the linear range of the recombinant ACE2-Fc protein as standard. After adjustment for molecular weight differences, the standard plasma had a concentration of 1.01 mg/ml anti-SARS-CoV-2 S IgG. Aliquots of this plasma sample were used as standards for quantification of seven randomly selected sera in the flow cytometric assay (Fig. 4B) and the EuroImmun ELISA (Fig. 4C), respectively. This revealed a good correlation for the two quantification methods (Fig. 4 D). Within PCR-positive individuals, the flow cytometric assay could monitor serum antibody responses to the SARS-CoV-2 S protein in the range of 10 μg/ml (mild cases) up to 6 mg/ml in severely sick patients. Thus, this quantification provides an objective value of SARS-CoV-2 spike-binding IgG concentrations in a given sample that can be compared among different assays and laboratories.

**Figure 4.**
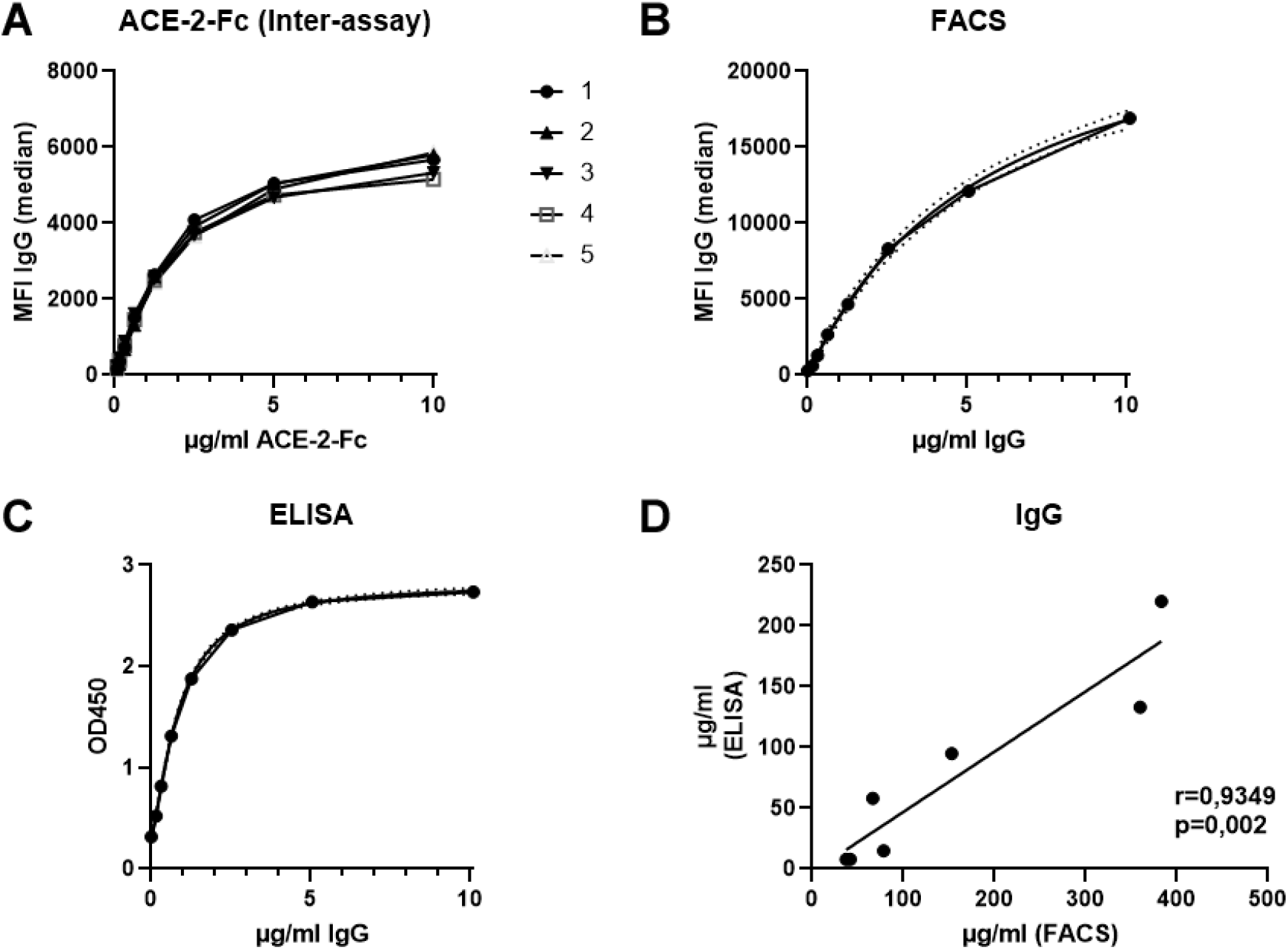
Quantification of SARS-CoV-2-specific antibody levels by an external ACE-2-Fc standard. (A) Defined concentrations of the ACE-2-Fc protein were analysed by the flow cytometric assay in five independent measurements (each symbol representing one measurement). With the help of this ACE-2-Fc standard, the concentration of a standard serum was defined. The standard serum was measured in a dilution series by the flow cytometric assay (B) and the EuroImmun ELISA (C). 4-PL curve fitting (shown with 95% confidence bands) was used to generate a standard curve for the absolute quantification of unknown samples. (D) Seven randomly chosen sera from PCR-confirmed COVID-19 patients were quantified by ELISA and the cytometric assay for SARS-CoV-2-specific IgG. The plot assesses the correlation between those two assays by spearman’s rank correlation coefficient.

## Discussion

While rigorous measures led to a partial control of the recent SARS-CoV-2 pandemic in some countries, validated serological assays are urgently needed to consolidate those achievements and to support the transition to a post-peak phase. This includes for example diagnostic measures for late/post infection stages, COVID-19 contact tracing, the assessment of epidemiological aspects, and the evaluation of immunity after infection or in potential vaccine trials. In the recent study, we validated an in-house flow cytometric assay for the detection of SARS-CoV-2 S-specific IgM and IgG using sera from PCR-confirmed COVID-19 cases and a collection of control serum samples. In regard to specificity and sensitivity, our flow cytometric assay showed a comparable or even better performance compared to commercial CE-marked serological assays (EuroImmun ELISA and Shenzhen Yhlo CLIA).

Detection of viral nucleic acids via real time PCR is the gold standard in the diagnosis of acute SARS-CoV-2 infections. However, despite its reliability early during infection, confirmation of an infection at later time points becomes less reliable. As early as eight days post-infection, the diagnostic value of serological assays might therefore outperform nucleic-acid-based methods (15,16). Indeed, also our study showed seroconversion in a longitudinal set of sera from one patient eight days after the first positive PCR test, although the exact infection date is not clearly defined. As reported before (15,17,18), IgM did not generally possess a higher clinical sensitivity compared to IgG, since most of the seropositive specimen tested in the present study were positive for both isotypes. Detection of SARS-CoV-2-specific IgA was previously reported as more sensitive than detection of IgG in the EuroImmun ELISA kits (19). However, while this held also true in our study, IgG and IgM measured by the flow cytometric assay were similarly sensitive compared to the IgA ELISA. This higher sensitivity to detect S-specific IgG might be due to the different viral antigens used in the assays. Our flow cytometric assay exploits full-length S protein in its natural conformation and with the respective posttranslational modifications due to the expression in mammalian cells. This enables detection of the full spectrum of S-specific antibodies directed against conformational epitopes and glycosylated sites as well, some immunogenic sites possibly missing in truncated, recombinant S1-only proteins as used in the EuroImmun ELISA.

A potential downside of using full-length S for serological testing might be the detection of cross-reactive antibodies induced by other HCoV. Along this line, some assays detect only antibodies directed against the S1 subunit (like the EuroImmun ELISA) or the receptor binding domain in order to increase specificity (19,20). However, in a collection of sera from individuals that suffered from an infection with an endemic HCoV shortly before blood collection, none was tested positive for SARS-CoV-2 antibodies. In additional 180 specimens sampled before the COVID-19 outbreak, two sera were found to be reactive in the flow cytometric assay. Since endemic HCoV seroprevalence is high in the general population (11) and those two individuals were non-reactive in the commercial S1-specific ELISA, a plausible explanation is cross-reaction of antibodies induced by the endemic HCoVs with the S2 subunit of SARS-CoV-2. Although the reactivity of the two specimens need to be classified as false-positive detection of SARS-CoV-2 antibodies, these cross-reactive antibodies might possess antiviral activity against COVID-19 and the analysis of cross-protection due to these responses might be an interesting topic for further investigations. Although a clinical specificity of about 99% seems high, the positive predictive value of commonly used antibody tests under the currently expected SARS-CoV-2 seroprevalence rates in many countries of 1-2% is low. While this may be controlled for in seroepidemiological studies, confirmatory tests are urgently needed if serological assays are to be used for individual diagnosis of past SARS-CoV-2 infections.

Regarding the clinical sensitivity, the flow cytometric serology assay detected 100% of IgM and IgG positive samples measured by either of the two commercial assays. Only in cases were blood samples were taken at the same day as PCR sampling, all assays (ELISA, CLIA, cytometry) were negative, probably reflecting acute infections prior to development of detectable antibody responses. The lower analytical detection limit of the flow cytometric assay is consistent with its excellent clinical sensitivity.

As the demand for SARS-CoV-2 serology testing kits increases worldwide and only a limited number of suppliers have developed such kits yet, there is a high need to expand the portfolio of serology techniques to meet this demand. The flow cytometry-based technique to detect SARS-CoV-2 seroconversion presented here, fulfils fundamental criteria in regard to sensitivity, specificity, and robustness. Therefore, we think that this method combines a moderate workload without the need for critical components and the possibility for high-throughput testing and, thus, can expand the existing portfolio. The basic requirements needed like cell culture, plastic ware, and a flow cytometer are available in many standard diagnostic and biomedical research labs. Thus, this method can increase serology testing capacities significantly without competing for ELISA/CLIA kits. Given the large number of antibody assays reaching the market without clearly defined analytical sensitivities, using recombinant ACE-2 Fc protein for standardization is a potential strategy for cross-assay comparisons. Moreover, the quantification of S-specific antibody responses might help to define protective antibody levels as correlate of protective immunity.

In conclusion, our in-house flow cytometry-based serological assay has good specificity and sensitivity for the detection of antibodies to SARS-CoV-2. In addition to the nucleotide sequence of the antigen, only readily available reagents were needed to establish the assay. Therefore, the flow cytometric assay may also serve as a rapid-response antibody test against other emerging viral infections.

## Data Availability

All relevant data are within the manuscript.

